# Medical treatment of 55 patients with COVID-19 from seven cities in northeast China who fully recovered: a single-center, retrospective, observational study

**DOI:** 10.1101/2020.03.28.20045955

**Authors:** Lichao Fan, Huan Liu, Na Li, Chang Liu, Ye Gu, Yongyu Liu, Yu Chen

## Abstract

**Background:** COVID-19 is an emerging disease caused by the SARS-CoV-2 virus; no specific medication has been identified to date. We aimed to investigate the administered medications and intervention times for patients who completely recovered from COVID-19.

**Methods:** This single-center, retrospective, and observational study included 55 patients with COVID-19 who were transferred to Shenyang Sixth People’s Hospital between January 20 and March 15, 2020. Demographic information, symptoms, laboratory indicators, treatment processes, and clinical outcomes were collected. Administered drugs and intervention times were compared in 47 and eight patients with mild and severe symptoms, respectively.

**Findings:** All 55 patients recovered. Fifty-three patients (96·36%) received antiviral therapy, including 45 in the mild group (median treatment: 14 days; 17 received umifenovir) and all eight severe-group patients (median treatment: 17·5 days; four received lopinavir/ritonavir). Twenty-nine patients (52·72%) were administered antibiotics, including 21 in the mild group (median treatment: 13·5 days; 15 received moxifloxacin) and all eight in the severe group (median treatment: 9 days; two received linezolid). Moreover, seven patients (12·72%) were treated with glucocorticoids and nine (16·36%) with immunomodulators.

**Interpretation:** Given the 100% recovery rate, early administration of antiviral drugs can be considered. Umifenovir may benefit patients with mild symptoms, while lopinavir/ritonavir may benefit those with severe symptoms. Prophylactic administration of common antibiotics may reduce the risk of co-infection. The use of glucocorticoids is usually not necessary.

**Funding:** This work was supported by the Shenyang Major Science and Technology Innovation R&D Program (JY2020-9-018 to Y. Chen).

## Introduction

The pathogenesis of SARS-CoV-2 infection remains unclear, and effective drugs and regimens for the treatment of COVID-19 have not been identified.^1-2^ In China, nationally recommended trial-based antiviral and other symptom-managing drugs are administered. As such, a retrospective review of the types and doses of clinical drugs, courses of treatment, and intervention times in patients cured of COVID-19 would be highly informative for treating patients with this disease worldwide.

Data on medications that were administered to patients who ultimately recovered from COVID-19 are scarce but crucial for clinicians. To that end, we investigated 55 patients confirmed to have COVID-19 who completely recovered after being transferred to Shenyang Sixth People’s Hospital, a designated treatment facility in Liaoning Province. Our study included data from patients with both mild and severe symptoms, and helped identify the drugs and administration methods required at different stages of the disease to help patients recover.

## Methods

### Study design and participants

We performed a single-center, retrospective, and observational study at Shenyang Sixth People’s Hospital (Shenyang, Liaoning, China), a government-designated centralized medical facility for the treatment of patients with COVID-19 in Liaoning Province. All patients were from hospitals that received patients for initial COVID-19 treatment in seven cities in Liaoning Province, including Shenyang Chest Hospital. According to the Interim Guidelines issued by the World Health Organization (WHO) on January 12, 2020,^3^ patient throat swabs and sputum samples were collected and subjected to a nucleic acid (RNA) test for COVID-19. We included all 55 consecutive patients with COVID-19 who were treated between January 20 and March 15, 2020; none were excluded. Patients were considered the ‘mild’ group (mild/moderate), whereas those were considered the ‘severe’ group (severe/critical); these classifications were according to the criteria stated in the COVID-19 Diagnosis and Treatment Plan issued by the National Health Commission of the People’s Republic of China.^2^

The study was reviewed and approved by the Ethics Committee of Shenyang Chest Hospital (approval number: KYXM-2020-001-01) and was also documented by the Ethics Committee of the Shenyang Sixth People’s Hospital. Written informed consent forms were waived owing to the rapid development of the infectious COVID-19 disease.

### Data collection

We reviewed clinical manifestations as well as laboratory and radiological findings of all enrolled patients and collected data that included age, sex, epidemiological history, past history, symptoms, complications, laboratory indicators, therapeutic drugs, and intervention time.

### Outcomes

The endpoint was the total patient recovery rate; individuals meeting the discharge criteria were included in the ‘recovered’ statistics. These discharge criteria were consistent with China’s COVID-19 Diagnosis and Treatment Plan^2^ as follows: 1. Body temperature returned to normal and remained so for at least 3 days; 2. respiratory symptoms were appreciably relieved; 3. pulmonary imaging showed a significant improvement in acute exudative lesions; and 4. nucleic acid tests of the sputum, nasopharyngeal swabs, and other respiratory specimens were negative twice consecutively following a minimum interval of 24 hours.

### Statistical analysis

Given that the purpose of this study was to examine the clinical characteristics and drug administration data for patients with COVID-19, no formal hypothesis was established with which to calculate the optimal sample size. Continuous variables are expressed as means (standard deviations) or medians (interquartile ranges [IQRs]), while categorical variables are denoted as percentages.

## Results

The mean age of the 55 patients in our study was 46·8 years. Among them, 30 (54·55%) were male, 28 (50·91%) had been in Wuhan/Hubei, and 19 (34·55%) were complicated with other chronic diseases. Lung computed tomography scans showed local or diffuse infiltration shadows in 54 patients (98·18%), whereas the remaining patient (1·82%) had no inflammatory changes. There were 47 patients (85·45%) in the mild group and eight (14·55%) in the severe group (Table 1). The most common symptoms of COVID-19 were fever 32 (58·18%) and cough 27 (49·09%). Seventeen patients (30·91%) were complicated with liver function impairment, 15 (27·27%) with hypoxemia, and two (3·64%) with acute respiratory distress syndrome (ARDS) (Table 2). The white blood cell counts, lymphocyte counts, and percentage of lymphocyte counts of patients in the mild group were in the normal range, although C-reactive protein levels (15·73 mg/L) were elevated. In the severe group, however, lymphocyte counts (0·78 × 10^9^ /L) and the percentage of lymphocytes (12·30%) were suppressed, while C-reactive protein levels (47·21 mg/L) were elevated (Table 2). In the mild and severe groups, the median durations for the lymphocyte counts to return to normal were 11 and 9 days, respectively; those for lung shadows to markedly improve were 12 and 19 days, respectively; and those for achieving negative COVID-19 RNA conversion were 12 and 19 days, respectively (Figure 1).

**Table 1.** Epidemiological and baseline characteristics of 55 patients with COVID-19.

| Clinical characteristics | Total (n = 55) | Mild group (n = 47) | Severe group (n = 8) |
| --- | --- | --- | --- |
| Age (years) | 46.8 | 46.40 | 46.12 |
| Sex (Male/Female) | 30/25 | 24/23 | 6/2 |
| Epidemiological history |  |  |  |
| Have been to Wuhan/Hubei | 28 (50.91%) | 22 (46.81%) | 6 (75.00%) |
| Have been in contact with confirmed cases | 18 (32.72%) | 16 (34.04%) | 2 (25.00%) |
| Cluster onset | 7 (12.72%) | 7 (14.89%) | 0 |
| Other | 2 (3.63%) | 2 (4.26%) | 0 |
| Preexisting medical conditions |  |  |  |
| Diabetes | 8 (14.55%) | 7 (14.89%) | 1 (12.50%) |
| Coronary artery disease | 3 (5.45%) | 2 (4.26%) | 1 (12.50) |
| Hypertension | 8 (14.55%) | 8 (17.02%) | 0 |

**Table 2.** Symptoms, complications, and laboratory test results of 55 patients with COVID-19.

| Clinical characteristics | Total (n = 55) | Mild group (n = 47) | Severe group (n = 8) |
| --- | --- | --- | --- |
| Symptoms and signs |  |  |  |
| Fever | 32 (58.18%) | 24 (51.06%) | 8 (100%) |
| Headache | 2 (3.64%) | 2 (4.26%) | 0 |
| Cough | 27 (49.09%) | 24 (51.06%) | 3 (37.50%) |
| Sore throat | 7 (12.73%) | 5 (10.64%) | 2 (25.00%) |
| Fatigue | 8 (14.55%) | 8 (17.02%) | 0 |
| Shortness of breath | 8 (14.55%) | 5 (10.64%) | 3 (37.50%) |
| Nausea and vomiting | 4 (7.27%) | 2 (4.26%) | 2 (25.00%) |
| Diarrhea | 6 (10.91%) | 4 (8.51%) | 2 (25.00%) |
| Muscle or joint pain | 7 (12.73%) | 5 (10.64%) | 2 (25.00%) |
| Complications |  |  |  |
| Acute respiratory distress syndrome | 2 (3.64%) | 0 | 2 (25.00%) |
| Hypoxemia | 15 (27.27%) | 7 (14.89%) | 8 (100.00%) |
| Liver dysfunction | 17 (30.91%) | 11 (23.40%) | 6 (75.00%) |
| Laboratory tests |  |  |  |
| White blood cell count ($\times 10^9/L$) | 6.13 | 5.78 | 8.18 |
| Lymphocyte count ($\times 10^9/L$) | 1.61 | 1.76 | 0.78 |
| Lymphocyte percentage (%) | 28.76 | 31.56 | 12.30 |
| C-reactive protein (mg/L) | 20.31 | 15.73 | 47.21 |
| D-dimer (mg/L) | 0.62 | 0.48 | 1.44 |
| Procalcitonin (ng/mL ) | 0.08 | 0.07 | 0.11 |
| Total bilirubin ($\mu\text{mol/L}$) | 19.47 | 18.98 | 22.38 |
| Alanine aminotransferase (U/L) | 40.65 | 37.85 | 57.13 |
| Creatinine ($\mu\text{mol/L}$) | 54.25 | 54.13 | 55.00 |

**Figure 1:**
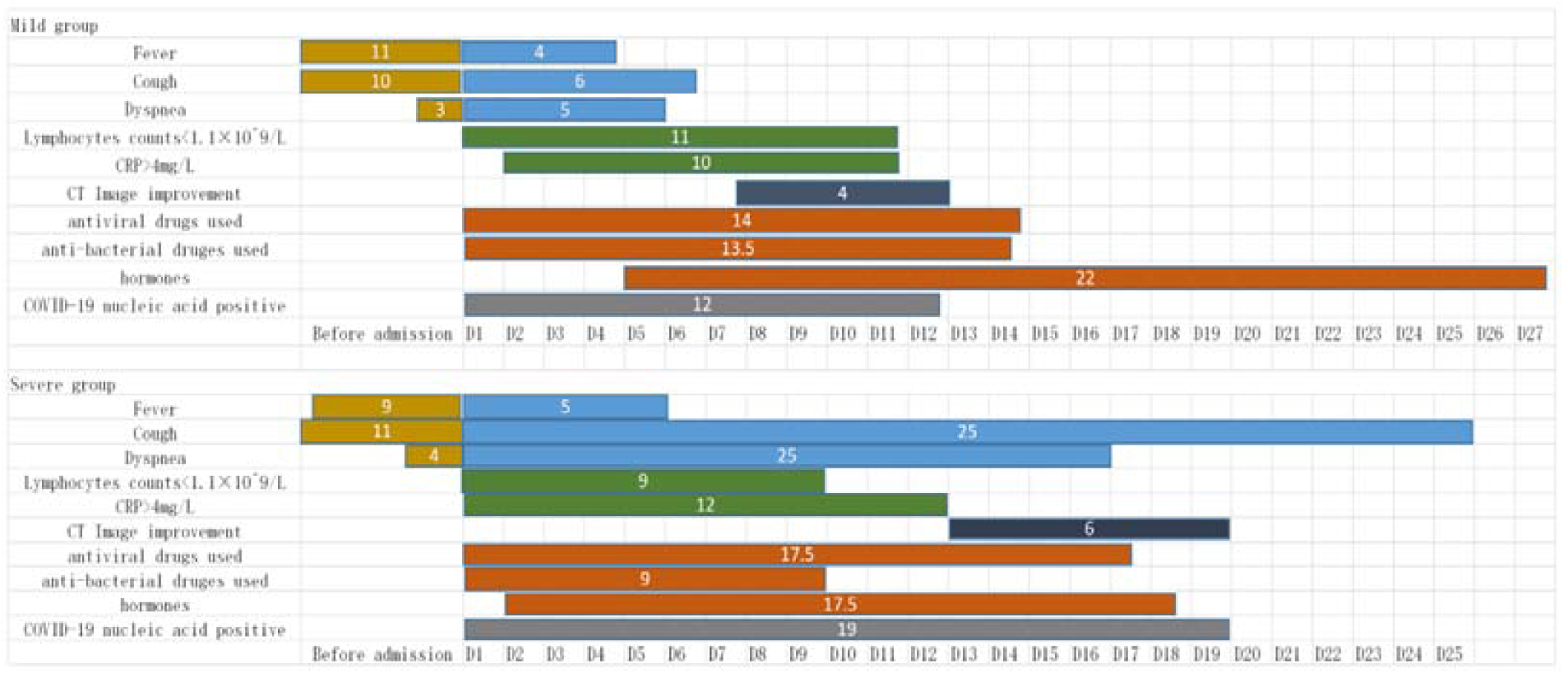
Clinical courses of major symptoms and medical treatment and duration of the viral shedding from illness onset in patients hospitalised with COVID 19. Figures show median duration of symptoms, abnormal laboratory indicators and medical treatment. COVID-19=coronavirus disease 2019,CRP=c-reactive protein,CT=Computed tomography,D=Days after illness onset.

Fifty-three patients (96·36%) received antiviral therapy for a median time of 14 days (IQR 12–18 days), while two patients (3·64%) were not administered antiviral drugs (one was a pregnant woman and the other had asymptomatic infection). Among those who received antiviral drugs, 45 were in the mild group (95·74% of this group); their median treatment time was 14 days (IQR 12–17 days) and 17 of them (37·78%) were treated with umifenovir, 17 (37·78%) with umifenovir + lopinavir/ritonavir, and five (11.11%) with lopinavir/ritonavir. Moreover, all eight patients in the severe group received antiviral drugs, with a median treatment time of 17·5 days (IQR 11–19·25 days). Four patients in the severe groups (50%) were treated with lopinavir/ritonavir, three (37·50%) with umifenovir + lopinavir/ritonavir, and one (12·50%) with umifenovir. Twenty-nine patients (52·72%) were treated with antibiotics for a median time of 10 days (IQR 8·5–15); 19 of these 29 patients (65·52%) were treated with moxifloxacin while three (10·34%) received linezolid. Among the patients treated with antibiotics, 21 were in the mild group (44·68% of this group); their median treatment time was 13·5 days (IQR 5·75–9·25 days), with 15 (71·42%) treated with moxifloxacin and two (9·52%) receiving carrimycin. The remaining antibiotic recipients comprised all eight patients in the severe group (100%), with a median treatment time of 9 days (IQR 9·75–15·25); four patients (50%) were treated with moxifloxacin and two (25%) with linezolid. Seven patients (12·72%) were treated with glucocorticoids, 20 (36·36%) received recombinant human interferon alpha-1b, and nine (16·36%) were treated with thymalfasin (Figures 1–3).

**Figure 2.**
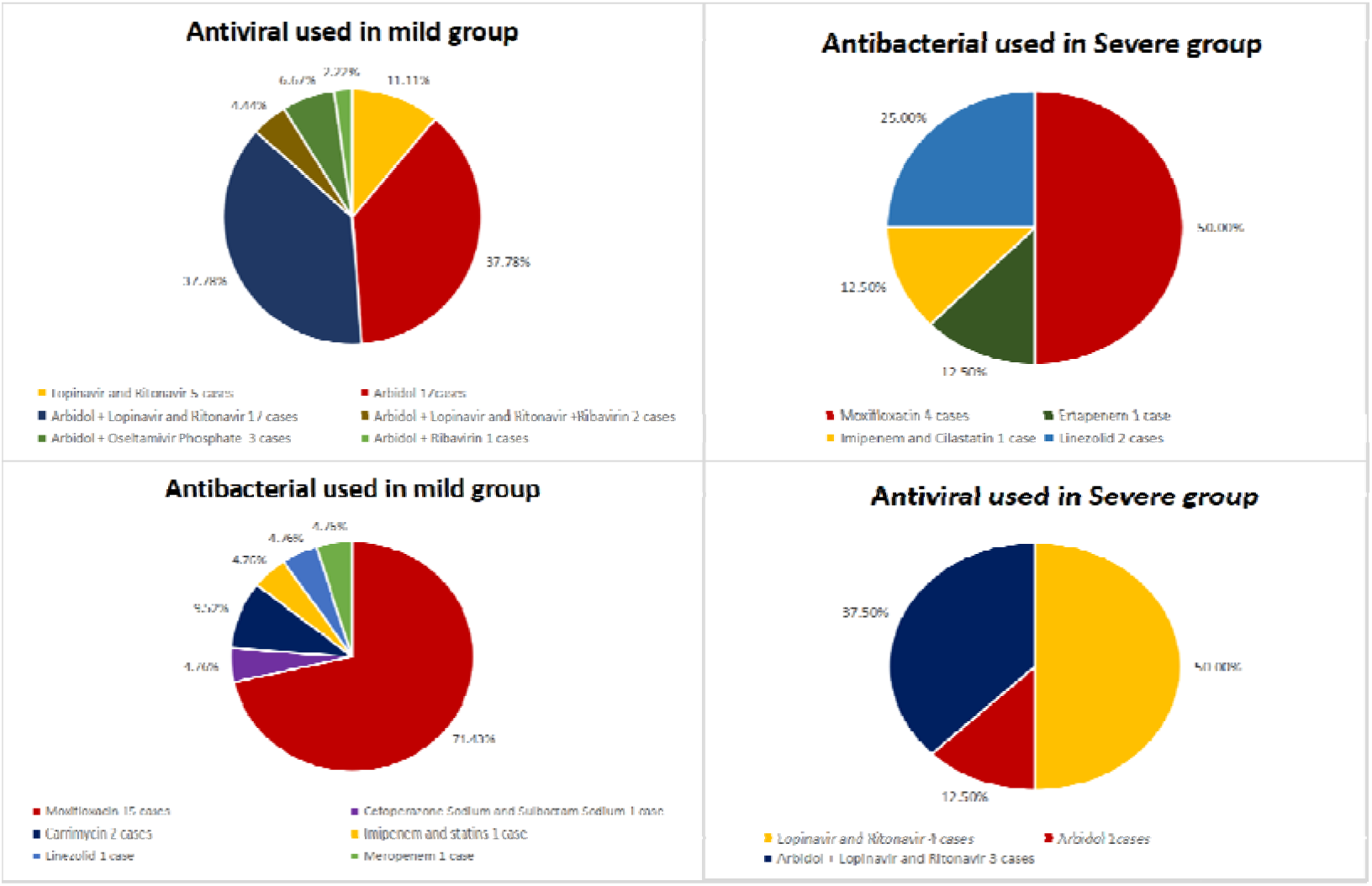
Use of antiviral drugs and antibiotics in mild and severe patients.

**Figure 3.**
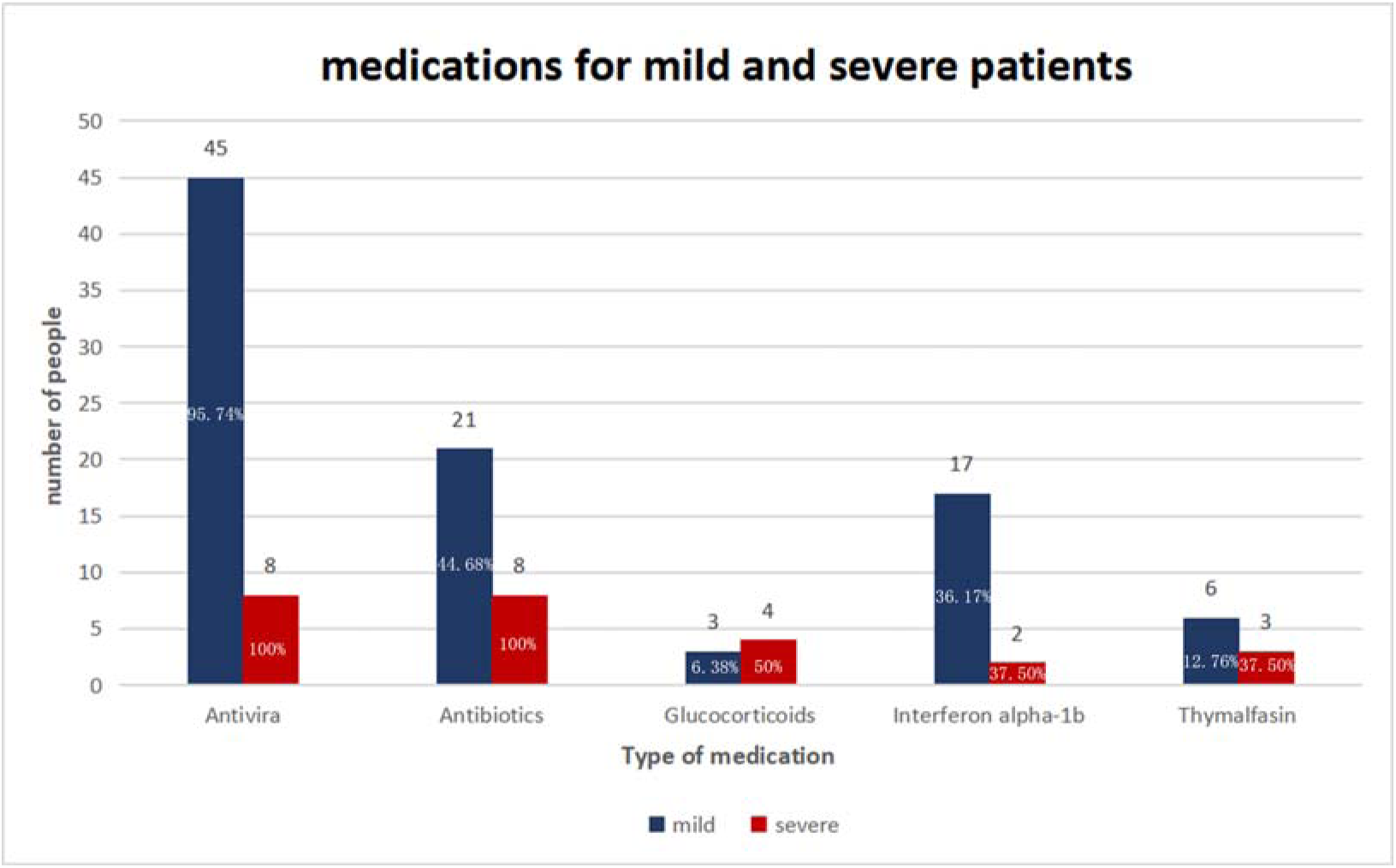
Medications for mild and severe patients.

## Discussion

Patients with COVID-19 in the present study achieved a 100% recovery rate. No effective antiviral drugs that treat COVID-19 have been identified to date, and opinions on whether antiviral drugs should even be used to treat COVID-19 differ.^1^ The National Health Commission of the People’s Republic of China has repeatedly issued and revised the COVID-19 Diagnosis and Treatment Plan, which recommends antiviral drugs such as lopinavir/ritonavir, ribavirin, umifenovir, and alpha-interferon.^2^ In the present study, 53 patients (96·36%) received antiviral therapy early in the course of their disease for a median time of 14 days (IQR 12–18 days). In the mild group, 17(37·78%) of the patients were treated with umifenovir and another 17 (37·78%) received a combination of umifenovir + lopinavir/ritonavir for a median time of 14 days (IQR 12–17 days). Liu et al.^4^ described the effectiveness of umifenovir in emerging respiratory infectious diseases such as influenza A (H1N1). Ji et al.^5^ also confirmed the efficacy of umifenovir for the treatment of coronavirus infection in an *in vitro* study. Our present findings indicated that umifenovir might benefit patients with mild symptoms. In the severe group, eight patients (100%) were administered antiviral drugs for a median time of 17·50 days (IQR 11–19·25 days); 4 (50%) were treated with lopinavir/ritonavir and 3 (37·50%) received a combination lopinavir/ritonavir regimen. Chu et al.^6^ found that lopinavir/ritonavir could inhibit coronavirus replication to some extent, thereby reducing the risk of ARDS or death in patients with severe acute respiratory syndrome (SARS). Chan et al.^7^ confirmed the efficacy of the combination of lopinavir/ritonavir and interferon-β for the treatment of MERS-CoV infection in animal models. In the present study, the use of lopinavir/ritonavir in the severe group was apparently effective in mitigating fever symptoms, promoting lung shadow absorption, and rapidly restoring the number of lymphocytes. The efficacy and safety of lopinavir/ritonavir are expected to be verified in future clinical randomized controlled trials. Ribavirin and interferon are also mentioned in the COVID-19 Diagnosis and Treatment Plan;^2^ in the present study, four patients in the mild group (7·27%) were administered ribavirin combined with antiviral therapy. Nucleoside analogs theoretically ought to possess anti-coronavirus activity to a certain extent;^8^ however, ribavirin was found to have a minimal antiviral effect against coronavirus *in vitro*.^9^ In the present study, 20 patients received aerosol inhalation of alpha-interferon soon after diagnosis (50 µg, twice per day). A retrospective study of patients with Middle East Respiratory Syndrome (MERS) in 2019 showed that interferon did not accelerate virus clearance,^10^ while a study of patients with SARS showed that alpha-interferon did not improve the patients’ prognosis.^11^ In the present study, a small number of patients were treated with ribavirin, and alpha-interferon was simply used to assist aerosol inhalation. Therefore, the usefulness of these two drugs for patients with COVID-19 is difficult to evaluate.

In terms of antimicrobial use, the WHO recommends empirical antimicrobial therapy based on the clinical diagnosis.^3^ China’s COVID-19 Diagnosis and Treatment Plan^2^ also emphasizes the avoidance of blind or inappropriate use of antibiotics. Kim et al.^12^ found that 38% of their patients with H1N1 infection developed secondary bacterial pneumonia 48 hours after admission to the intensive care unit, and that early empirical treatment helped improve their prognosis. Experience with SARS^13^ and MERS^14^ also suggests that prophylactic antibiotics may be appropriate after assessing the risk of co-infection in patients with severe symptoms.

Bacterial infection rates after COVID-19 infection remain unclear. In the present study, 12 (57·14%) of patients in the mild group were administered prophylactic drugs and 9 (42·86%) underwent empirical treatment (mainly with single-antibiotic moxifloxacin); the possibility of atypical pathogenic bacteria such as *Mycoplasma pneumoniae* infection was also considered. In the severe group, 5 (62·50%) of patients were administered prophylactic medication and 3 (37·50%) received empirical treatment. Prophylactic medication was administered to patients complicated with diabetes, chronic lung diseases, and ARDS who were at the early stage of receiving glucocorticoids. Broad-spectrum antimicrobials were recommended for patients at a higher risk of multidrug-resistant bacterial infections in the severe group, with drug doses adjustable according to the results of pathogen cultures. No evidence of secondary bacterial infection was observed in patients who received prophylactic medication. The present study suggested that the early and prudent use of prophylactic antibiotics in patients with COVID-19 may help reduce the risk of co-bacterial infections.

The WHO does not recommend the systematic use of glucocorticoids for viral pneumonia or concurrent ARDS.^3^ China’s COVID-19 Diagnosis and Treatment Plan recommends hormones as adjuvant therapy.^2^ In the present study, 48 (87·28%) of the patients did not receive glucocorticoids, while 7 (12·72%) did receive such agents during the rapid progression of their disease to inhibit inflammation and improve oxygenation at a dose of 1–2 mg/kg/day. Treatment was gradually reduced over 5–7 days until discontinuation, and showed no adverse reactions. As such, glucocorticoids appear to be unnecessary for patients with mild manifestations of COVID-19, while their use in treating patients with severe disease is controversial.

China’s COVID-19 Diagnosis and Treatment Plan^2^ suggests that immunotherapy can be attempted for patients with severe COVID-19. In the present study, 9 (16·36%) of the patients were administered thymalfasin. There has been no additional evidence gathered regarding the effectiveness of immunomodulatory drugs for viral pneumonia;^15^ hence, this notion requires further clinical observation and study. While high-flow oxygen therapy, invasive mechanical ventilation, and extracorporeal membrane oxygenation were provided to patients in the present study, their use was not investigated.

Our findings suggest that, while specific antiviral drugs are yet to be developed, currently available antiviral agents should be considered when treating patients with COVID-19. The prophylactic administration of single antiviral drugs to patients with severe symptoms, as well as to a proportion of those with mild manifestations, may help reduce the risk of co-infection. However, the use of glucocorticoids and immunomodulators needs further study.

Our study had some limitations given its single-center, retrospective, and observational nature. Owing to its small sample size, only descriptive data were available, and no statistical analyses were performed. Hence, randomized, double-blind, and controlled trials remain necessary for more accurate conclusions. Nevertheless, our data ought to provide helpful preliminary information at this stage of the COVID-19 pandemic.

## Data Availability

The data used to support the findings of this study are available from the corresponding author upon request.

## Contributors

Yu Chen and Yongyu Liu design the study. Chang Liu and Na Li responsible for literature search. Lichao Fan and Ye Gu collected the epidemiological and clinical data. Huan Liu and Lichao Fan processed statistical data, Huan Liu and Yu Chen drafted the manuscript. Yongyu Liu revised the final manuscript.

## Acknowledgments

This study was funded by the Shenyang Major Science and Technology Innovation R&D Program (JY2020-9-018 to Y. Chen). We thank all patients involved in the study. We would like to thank Editage (www.editage.cn) for English language editing

## References

1. Liu C, Zhou Q, Li Y, et al. Research and development on therapeutic agents and vaccines for COVID-19 and related human coronavirus diseases. ACS Cent Sci 2020; doi: 10.1021/acscentsci.0c00272.

2. National Health Commission of the PRC. COVID-19 diagnosis and treatment plan (trial version 5). Zhongguo Zhong Xi Yi Jie He Za Zhi 2020; doi: 10.7661/j.cjim.20200202.064.

3. World Health Organization. Clinical management of severe acute respiratory infection when novel coronavirus (2019-nCoV) infection is suspected: interim guidance. Jan 28, 2020. Available at: https://www.who.int/publications-detail/clinical-management-of-severe-acute-respiratory-infection-when-novel-coronavirus-(ncov)-infection-is-suspected (accessed Mar 24, 2020).

4. Liu Q, Xiong HR, Lu L, et al. Antiviral and anti-inflammatory activity of arbidol hydrochloride in influenza A (H1N1) virus infection. Acta Pharmacologica Sinica 2013; 34: 1075–83.

5. Ji X, Zhao Y, Zhang M, Zhao J, Wang J. In vitro experimental study on the effect of Arbidol against SARS virus. Pharmaceutical Journal of Chinese PLA 2004; 20: 274–6.

6. Chu MC, Cheng VC, Hung IF, et al. Role of lopinavir/ritonavir in the treatment of SARS: initial virological and clinical findings. Thorax 2004; 59: 252–6.

7. Chan JF, Yao Y, Yeung ML, et al. Treatment with lopinavir/ritonavir or interferon-β1b improves outcome of MERS-CoV infection in a non-human primate model of common marmoset. J Infect Dis 2015; 212: 1904–10.

8. Minskaia E, Hertzig T, Gorbalenya AE, et al. Discovery of an RNA virus 3’->5’ exoribonuclease that is critically involved in coronavirus RNA synthesis. Proc Natl Acad Sci U S A 2006; 103: 5108–113.

9. Smith EC, Blanc H, Surdel MC, Vignuzzi M, Denison MR. Coronaviruses lacking exoribonuclease activity are susceptible to lethal mutagenesis: Evidence for proofreading and potential therapeutics. PloS Pathog 2013; 9: e1003565.

10. Arabi YM, Shalhoub S, Mandourah Y, et al. Ribavirin and interferon therapy for critically ill patients with Middle East respiratory syndrome: a multicenter observational study. Clin Infect Dis 2019; doi: 10.1093/cid/ciz544.

11. Stockman LJ, Bellamy R, Garner P. SARS: Systematic review of treatment effects. PLoS Med 2006; 3: e343.

12. Kim SH, Hong SB, Yun SC, et al. Corticosteroid treatment in critically ill patients with pandemic influenza A/H1N1 2009 infection: analytic strategy using propensity scores. Am J Respir Crit Care Med 2011; 183: 1207–14.

13. Zhong NS, Zeng GQ. Our strategies for fighting severe acute respiratory syndrome (SARS). Am J Respir Crit Care Med 2003; 168: 7–9.

14. National Health Commission of the PRC. MERS Diagnosis and Treatment Plan (2015). Zhongguo Bing Du Bing Za Zhi 2015; 5: 352–4.

15. Liu WJ, Zhao M, Liu K, et al. T-cell immunity of SARS-CoV: Implications for vaccine development against MERS-CoV. Antiviral Res 2017; 137: 82–92.

